# *HIPK4* is a novel gene associated with teratozoospermia and male infertility

**DOI:** 10.64898/2026.03.04.26346694

**Authors:** Sophie Adina Koser, Cynthia Rieck, Isabella Aprea, Claudia Krallmann, Avinash Satish Gaikwad, Julia Wallmeier, Retno Tenardi-Wenge, Sara Di Persio, Nina Neuhaus, Johanna Raidt, Heymut Omran, Sandra Laurentino, Sabine Kliesch, Birgit Stallmeyer, Corinna Friedrich, Frank Tüttelmann

## Abstract

**STUDY QUESTION:** Are pathogenic variants in *Homeodomain-interacting protein kinase* (*HIPK4*) associated with sperm head abnormalities causing male infertility?

**SUMMARY ANSWER:** *HIPK4* is a novel candidate gene associated with sperm head defects and human male infertility.

**WHAT IS KNOWN ALREADY:** Numerous genes causing male infertility due to Multiple Morphological Abnormalities of the sperm flagella (MMAF) have been described but the genetic basis of sperm head defects is less well understood.

**STUDY DESIGN, SIZE, DURATION:** Four infertile brothers displaying varying degrees of quantitatively and/or qualitatively impaired spermatogenesis, their parents, and their fertile brother were included in the study. Further, the Male Reproductive Genomics (MERGE) cohort comprising exome/genome sequencing data of >3,300 men was queried.

**PARTICIPANTS/MATERIALS, SETTING, METHODS:** We performed exome sequencing in all five brothers and their parents. To characterise the sperm phenotype, standard semen analysis, immunofluorescence staining, and transmission-electron microscopy (TEM) were carried out. Further, we evaluated the impact of the *HIPK4* variant in cell culture experiments using HEK293T cells.

**MAIN RESULTS AND THE ROLE OF CHANCE:** Analysing the exome data, we could not identify a common genetic cause in all four affected brothers. However, one of the affected brothers was compound heterozygous for two loss-of-function variants in *DNAH17* (c.1076_1077dup p.(Lys360*) and c.7752+2T>A p.?) associated with markedly reduced sperm motility and MMAF. The variants’ pathogenicity was further validated by TEM of flagellar cross-sections revealing an outer dynein arm defect and axonemal disruption. On the contrary, his three infertile brothers were homozygous for the start-loss variant c.1A>G in *HIPK4*. This gene is expressed during spermiogenesis and is reportedly involved in sperm head shaping in mice. Heterologous expression of (partial) *HIPK4* variant cDNA elucidated the alternative use of an in frame start codon located 35 amino acids downstream, resulting in an N-terminally truncated protein p.(Met1_Glu35del). The truncated HIPK4 protein lacks parts of its kinase domain and shows reduced protein stability. In line with published mouse models, all three brothers displayed 100% abnormal sperm head morphology with variable defects. Importantly, one brother affected by *HIPK4* variants fathered a child after successful intracytoplasmic sperm injection demonstrating that it is a treatment option for HIPK4-related teratozoospermia. No further men from the MERGE cohort were affected by biallelic *HIPK4* variants. Taken together, *HIPK4* is an autosomal-recessive candidate gene associated with sperm head defects and male infertility.

**LARGE SCALE DATA:** The reported variants in *DNAH17* and *HIPK4* were submitted to ClinVar.

**LIMITATIONS, REASONS FOR CAUTION:** Independent replication is required to assess the phenotypic spectrum and the reproductive outcome associated with biallelic *HIPK4* variants and to formally establish the gene-disease relationship for male infertility.

**WIDER IMPLICATIONS OF THE FINDINGS:** This study raises awareness of the significant genetic heterogeneity of male infertility. The described family highlights that distinct genetic causes may underlie a seemingly similar phenotype. Exome sequencing of families is helpful to efficiently disentangle individual causes among affected family members.

**STUDY FUNDING/COMPETING INTEREST(S):** N.N., J.R., H.O., S.L., C.F., and F.T. were supported by the Deutsche Forschungsgemeinschaft (DFG, German Research Foundation) within the Clinical Research Unit ‘Male Germ Cells’ (CRU326, project number 329621271). R.T.W., N.N., J.R., H.O., and F.T. were supported by the Federal Ministry of Research, Technology and Space (BMFTR) as part of the project ReproTrack.MS (grant 01GR2303). S.A.K. was supported by the DFG Clinician Scientist programme CareerS Münster (project number 493624047). A.S.G. was supported by the Medical Faculty Münster via an Innovative Medical Research (IMF) grant (GA-122104).

## Introduction

Sperm head abnormalities and other morphological defects (teratozoospermia) are frequently observed in men in infertile couples (Tüttelmann *et al*., 2018). Still, more than 70% of men do not receive a causal genetic diagnosis (Oud *et al*., 2025). While the number of genes associated with sperm flagellar defects has steadily grown during the past years, the genetic basis of sperm head abnormalities is less well understood (Arora *et al*., 2024; Beurois *et al*., 2020; Cavarocchi *et al*., 2025).

Disturbances of spermatogenesis, the complex process of male gametogenesis, can explain abnormal semen parameters. In order to understand the genetic causes of morphological sperm defects, it is necessary to focus on the processes of post-meiotic differentiation that occur during spermiogenesis – the drastic morphological transformation of haploid round spermatids into elongated spermatids and finally into sperm consisting of head, midpiece and tail.

Within the group of qualitative spermatogenic failure the term Multiple Morphological Abnormalities of the sperm Flagella (MMAF) describes combined defects with absent, short, bent, coiled flagella, and/or irregular calibre and is associated with severely reduced sperm motility (Touré *et al*., 2021). Considering MMAF, more than 20 genes have reached a sufficient level of evidence for their association to this phenotype, i.e., an at least moderate gene-disease relationship (GDR), and are ready for clinical diagnostic analyses (Stallmeyer *et al*., 2025).

MMAF can be caused by variants in genes encoding axonemal proteins such as subunits of the flagellar outer or inner dynein arms (ODAs/IDAs). One example is the ODA subunit dynein axonemal heavy chain 17 (DNAH17) (Milisav and Affara, 1998). Biallelic pathogenic variants in *DNAH17* cause impaired sperm motility (asthenozoospermia) and MMAF due to lack of ODAs and/or disorganisation of the axonemal ultrastructure of the sperm flagellum (Song *et al*., 2023; Whitfield *et al*., 2019).

Concerning sperm head defects, however, only a handful of genes are ready to be included in diagnostic gene panels (Stallmeyer *et al*., 2025). These few genes are associated with specific, monomorphic sperm head phenotypes such as macrozoospermia (*AURKC*), globozoospermia (*DPY19L2*), or acephalic sperm (*PMFBP1*, *SUN5*, *TSGA10*) (Stallmeyer *et al*., 2025).

Spermiogenesis including sperm head shaping requires fine-tuned regulation. Because spermatids are transcriptionally inactive, posttranslational regulation such as phosphorylation plays an important role (Baker, 2016; Li *et al*., 2019). Here, protein kinases come into play: they use ATP to phosphorylate their targets at specific residues and can thereby (in)activate protein functions (Röhm *et al*., 2021). One member, the serine/threonine kinase called homeodomain-interacting protein kinase (HIPK4; Arai *et al*., 2007; He *et al*., 2010) is reportedly required for sperm head shaping via cytoskeletal remodelling and male fertility in mice (Crapster *et al*., 2020; Liu *et al*., 2022). In humans, previously published *HIPK4* variants have been identified in men with azoospermia, either in a heterozygous state (Liu *et al*., 2022) or in a homozygous state but classified as a variant of uncertain significance (Alhathal *et al*., 2020). Accordingly, the relevance of *HIPK4* in human male infertility remains unsolved.

In this study, we present detailed genetic and phenotypic data of four brothers affected by infertility. Based on the morphological characterisation of sperm morphology and *in vitro* experiments, we disentangled two distinct genetic causes, variants in *DNAH17* and *HIPK4*, and thereby now describe a homozygous truncating variant in *HIPK4* explaining sperm head defects.

## Material/Subjects and Methods

### Subjects

Because of infertility, four brothers (M865, M1344, M1670, and M1611) from a Caucasian family presented at the Centre of Reproductive Medicine and Andrology (CeRA), University Hospital Münster between 2013 and 2018. According to their self-declaration, their parents were not related. Andrological examination included a detailed anamnesis, physical examination, testicular ultrasound, hormonal analysis (follicle-stimulating hormone [FSH], luteinising hormone [LH], testosterone [T]), and semen analysis. Routine genetic diagnostics ruled out chromosomal aberrations in all described men and AZF deletions in those men with sperm counts < 5 ×10^6^. Subsequently, the fertile brother (M1688) was recruited for clinical and genetic diagnostics and DNA samples were obtained from the parents for segregation analysis.

Further, the Male Reproductive Genomics (MERGE) cohort was queried for additional men affected by biallelic *HIPK4* variants. MERGE currently includes exome/genome sequencing data of n = 3,339 men in infertile couples, of whom 96 have normozoospermia and most have varying degrees of quantitatively and/or qualitatively impaired spermatogenesis (n = 3,243). The majority of subjects has crypto- or azoospermia (n = 577 and 2,038, respectively) and the remainder of 628 has oligo-, astheno- and/or teratozoospermia. All individuals provided written informed consent and the study was approved by the Ethics Committee of the Medical Faculty Münster (2010-578-f-S) in accordance with the Helsinki Declaration of 1975.

### Semen analysis

Repeated routine semen analyses were performed at the CeRA according to the WHO guidelines valid at the respective time (WHO, 2010; WHO, 2021). Importantly, strict criteria were always used when assessing sperm morphology after modified Papanicolaou staining. For visualisation of sperm morphology, pictures were taken with a PreciPoint O8 scanning microscope system with an 100x objective and oil.

### Exome/genome sequencing

Genomic DNA extracted from peripheral blood leukocytes was used for exome and genome sequencing as described before (Rotte *et al*., 2025). For exome sequencing, sample/library preparation and enrichment was done according to the protocol of Twist Bioscience’s Human Core Exome 1.3 plus RefSeq spike-ins or Human Core Exome 2.0 plus Comprehensive kit or Agilent’s SureSelect human all exon kits V4, V5, and V6. Genome samples were prepared using Illumina’s DNA PCR-Free library kit. For multiplexed sequencing, the libraries were index tagged using appropriate pairs of index primers. Quantity and quality of the libraries were determined with the ThermoFisher Qubit, the Agilent TapeStation 2200 or 4200, and the Tecan Infinite plate reader, respectively. Sequencing was performed on Illumina’s NextSeq 500, 550, 2000 or NovaSeq 6000 or X Plus systems using the corresponding reagent kits. Calling of single nucleotide variants (SNVs), small indels, and copy number variants (CNVs) was conducted using Broad’s GATK v3.8 or Illumina’s Dragen Bio-IT Platform v3.10, v4.3.6 or 4.3.13 using the reference genome version GRCh37.p13. The identified variants were then annotated using the Ensembl Variant Effect Predictor.

### Variant filtering

Variants were filtered for rare (minor allele frequency [MAF] < 0.01, gnomAD database v 2.1.1 (Karczewski *et al*., 2020), and < 0.01 in our in-house database), coding variants. Only high-impact variants (frameshift, premature stop codon, loss of start codon, splice acceptor/donor variants, and missense variants with a CADD score (Kircher *et al*., 2014) ≥ 20) were further assessed. After analysing diagnostic genes associated with qualitative spermatogenic failure (astheno-, teratozoospermia and others or mixed phenotypes) (Stallmeyer *et al*., 2025) for causal sequence or copy number variants, we performed an exome-wide analysis and focused on homozygous, (compound) heterozygous, and hemizygous sequence variants shared by the affected brothers. We only considered high-impact variants (see above) in genes expressed in the testis and excluded genes with a broad expression pattern via visual inspection of available RNA expression data (GTEx Consortium, 2020). Shared heterozygous and hemizygous variants were further inspected if their MAF was < 0.001 and if they were absent in the father and the fertile brother (M1688). Likewise, biallelic variants identified in the fertile brother were excluded. The prioritised variants in *HIPK4* and *DNAH17* were validated by Sanger sequencing (for primer sequences see Supplementary Table S1). Subsequently, the MERGE cohort was screened for potentially biallelic variants in *HIPK4* using the same filter criteria as described above. Regions of homozygosity were visualised using the AltAF plotter (Radtke *et al*., 2024).

### High-resolution immunofluorescence (IF) microscopy analysis

Semen was diluted to 1 mill./mL in Phosphate Buffered Saline (PBS) and spread onto glass slides for IF staining as described previously in (Aprea *et al*., 2021). In cases of severe oligozoospermia, where dilution to 1 mill./mL was not possible, samples were applied undiluted onto glass slides. After storage at −80 °C, cells were fixed with 4% paraformaldehyde, permeabilised with 0.1% Triton-X in PBS and blocked 2-4 h in 5% bovine serum albumin (BSA) in 0.1% Triton-X in PBS. To stain the flagellum, sperm were incubated overnight (4 °C) with a monoclonal mouse antibody against acetylated alpha-tubulin followed by incubation with the secondary antibody for one hour at room temperature in the dark (for antibody information see Supplementary Table S2). Nuclei were stained with Hoechst 33342 (1:1,000, Sigma, USA). Slides were imaged with a Laser Scanning Microscope (LSM 880, Carl Zeiss Microscopy GmbH, Germany), processed, and images were exported using the ZEISS ZEN Imaging Software 2012 (Carl Zeiss Microscopy GmbH, Germany). Figure panels were created with OMERO (Allan *et al*., 2012).

### Transmission electron microscopy (TEM)

TEM of sperm was performed as previously described (Aprea *et al*., 2023). Briefly, sperm were fixed in 2.5% glutaraldehyde over night at 4 °C. After pelleting and washing with tap water, samples were incubated at room temperature for 1.5–2 h in 1% osmium tetroxide. Following dehydration in an ethanol series, samples were first transferred to 1,2-epoxypropan and then incubated in a 1,2-epoxypropan-epon mixture (1:2) at 4 °C overnight. Finally, samples were embedded in epon and dried at 55 °C. Ultrathin sections (80 nm) of samples were placed on support grids and contrasted with 8% uranyl acetate. The samples were analysed with a transmission electron microscope Philips CM10 and TEM images acquired with a Quemesa camera and the iTEM SIS image acquisition software (both from Olympus Soft Imaging Solutions).

### Gene and protein expression and characterisation

*HIPK4* expression in human testicular tissues was evaluated in previously published bulk RNA sequencing data (Siebert-Kuss *et al*., 2023; European Genome-Phenome Archive: EGAD00001008652). Expression levels of *HIPK4* at single cell level were evaluated in human testis with normal spermatogenesis also using published datasets (Di Persio *et al*., 2021; NIH Gene Expression Omnibus (GEO): GSE153947). Expression levels were displayed as bubble plots using the ‘*DotPlot*’ function and as feature plots using the function ‘*FeaturePlot*’ in the Seurat R package (version 5.3.0).

The genomic sequence of *HIPK4* (ENST00000291823.3) was accessed via Ensembl (version 115) (Dyer *et al*., 2025). Amino acid (aa) alignments for different species and functional annotations (domains, features) of human HIPK4 or DNAH17 were retrieved from UniProt (The UniProt Consortium, 2023; see Supplementary Table S3 for the accession numbers). The common ATP-binding motif of kinase domains was added to the alignment (Röhm *et al*., 2021). The HIPK4 AlphaFold model (Jumper *et al*., 2021) was used for 3D visualisation of relevant residues of HIPK4 using ChimeraX (version 1.9) (Goddard *et al*., 2018).

### Cloning and mutagenesis

*HIPK4* exon 1 was amplified from genomic DNA derived from one patient homozygous for *HIPK4* c.1A>G and control DNA using PrimeSTAR Max polymerase (Takara Bio, Japan), cloned into the pcDNA3.1(+) vector (Thermo Fisher, USA) and modified by adding a C-terminal HA-tag and a stop codon. Human adult testis RNA (BioCat, Heidelberg, Germany) was used to clone the complete *HIPK4* cDNA. Reverse transcription was performed with the ProtoScript® II First Strand cDNA Synthesis Kit (New England Biolabs, USA). The QuikChange XL Site-Directed Mutagenesis Kit (Agilent, USA) was used to introduce the variant c.1A>G into the latter pcDNA3.1(+) construct. All primer sequences can be found in Supplementary Table S1. The correct insertion into the vector was confirmed by sequencing.

### Transfection of HEK293T cells

Human Embryonic Kidney cells (HEK293T, Leibniz Institute DSMZ-German Collection of Microorganisms and Cell Cultures, Germany) were transfected with 2 µg of the respective construct using the K2 Transfection System (Biontex, Germany) followed by medium exchange (Dulbecco’s Modified Eagle Medium containing 10% foetal bovine serum, Thermo Fisher, USA) after 6 h. 24 h (short construct) after transfection, cells were detached with ice-cold Dulbecco’s PBS (Thermo Fisher, USA), centrifugated (5 min, 4 °C, 300 x g), and resuspended with lysis buffer (100 mM NaCl, 20 mM Imidazol, 1% Triton X-100, 2 mM CaCl) containing a protease inhibitor cocktail (cOmplete™, Merck, Germany). After incubation on ice for 15 minutes and centrifugation (15 min, 4 °C, 15000 x g), the supernatant was stored at −20 °C or directly used for Western blot.

### Western blot

Cell lysates were diluted with Laemmli buffer (BioRad, USA) with dithiothreitol (DTT, Merck, Germany), denatured at 95 °C for 10 minutes and separated on a 12% acrylamide gel (short construct) or a 4-15% gradient gel (Mini-PROTEAN TGX Stain-free gels, Biorad, USA) (full protein). After blotting onto a PVDF membrane (Trans-Blot Turbo Transfer Pack, Biorad, USA) and blocking with 5% milk powder (AppliChem, Germany) in TBST, the membrane was incubated with the primary antibody (Supplementary Table S2) at 4 °C overnight. After incubation with HRP-conjugated secondary antibodies (Supplementary Table S2), chemiluminescence was detected with Peroxidase/Luminol system (Clarity enhanced chemiluminescence substrate, Biorad, USA) using a ChemiDoc MP Imaging System (Biorad, USA).

### Cycloheximide Chase Assay

HEK293T cells seeded in 6-well plates were transfected with 2 µg of the respective vector containing full-length *HIPK4* cDNA as described above. 48 h after transfection one well per condition was treated with 2 mL medium containing 100 µg/mL cycloheximide (Carl Roth, Germany) and further incubated for 24 h. Lysates of untreated (0 h) and treated cells (24 h) were used for Western blots with loading of equal volumes (10 µl). The membranes were cut horizontally to separate anti-HA and anti-GAPDH staining (Supplementary Table S2). Images were captured as described above and intensity of the bands was analysed using Fiji (ImageJ 1.54f) software. Protein levels were measured as mean grey values within a constantly sized rectangular selection in Western blots. The experiment was carried out four times independently (biological repeats) with averaging of technical repeats prior to statistical analysis.

### Statistical analyses

GraphPad Prism (version 10.2.3) was used for visualisation and statistical analysis of the cycloheximide assay. Data is presented as HIPK4 relative to GAPDH levels (HIPK4-HA/GAPDH). Normal distribution was confirmed via visual inspection of the QQ-plot. The proportion of residual protein after 24 h of incubation with cycloheximide [(HIPK4-HA/GAPDH)_24h_/(HIPK4-HA/GAPDH)_0h_] was compared between the wild type (WT) and mutant sample using a paired t-test with a significance level of p < 0.05 (one-tailed).

Plots showing expression of *HIPK4* in bulk RNA-sequencing and single cell RNA-sequencing data were generated with R (version 4.4.3) in the RStudio environment (version 2024.9.1.394).

### Variant classification and gene-disease relationship assessment

Variants were classified according to the guidelines of the American College of Medical Genetics and Genomics and the Association for Molecular Pathology (ACMG-AMP, ACMG used throughout the text) (Richards *et al*., 2015; Tavtigian *et al*., 2020) as described in Supplementary Table S4. A preliminary assessment of the gene-disease relationship was performed for *HIPK4* and male infertility according to ClinGen (Strande *et al*., 2017).

## Results

### Biallelic variants in DNAH17 and HIPK4 represent distinct causes of infertility within the family

To evaluate and compare phenotypes and genotypes among four brothers with infertility (M865, M1344, M1670, M1611), their parents, and an unaffected brother (M1688, Fig. 1), we performed an exome-based family analysis. Repeated semen analyses showed varying degrees of quantitatively and qualitatively impaired spermatogenesis in the affected brothers resulting in different combinations of oligo-, astheno-, and teratozoospermia (Human Phenotype Ontology [HPO] terms: HP:0000798, HP:0012207, HP:0012864). The three younger brothers M1344, M1760, and M1611 constantly displayed teratozoospermia with 100% sperm head defects. In contrast, the older brother M865 had 1 to 4% of sperm with normal morphology in more than 10 semen analyses (Fig. 1, Supplementary Table S5). Clinical history and examination, testicular ultrasound, as well as hormonal analysis (Supplementary Table S5) were unremarkable except for left-sided ligation of varicocele in M865. M865 and M1344 underwent successful ICSI and fathered three and one children, respectively, whereas their normozoospermic brother (M1688) spontaneously fathered three children. The percentage of regions of homozygosity throughout the exomes (Supplementary Fig. S1) indicated a distant familial relation of the parents.

**Figure 1.**
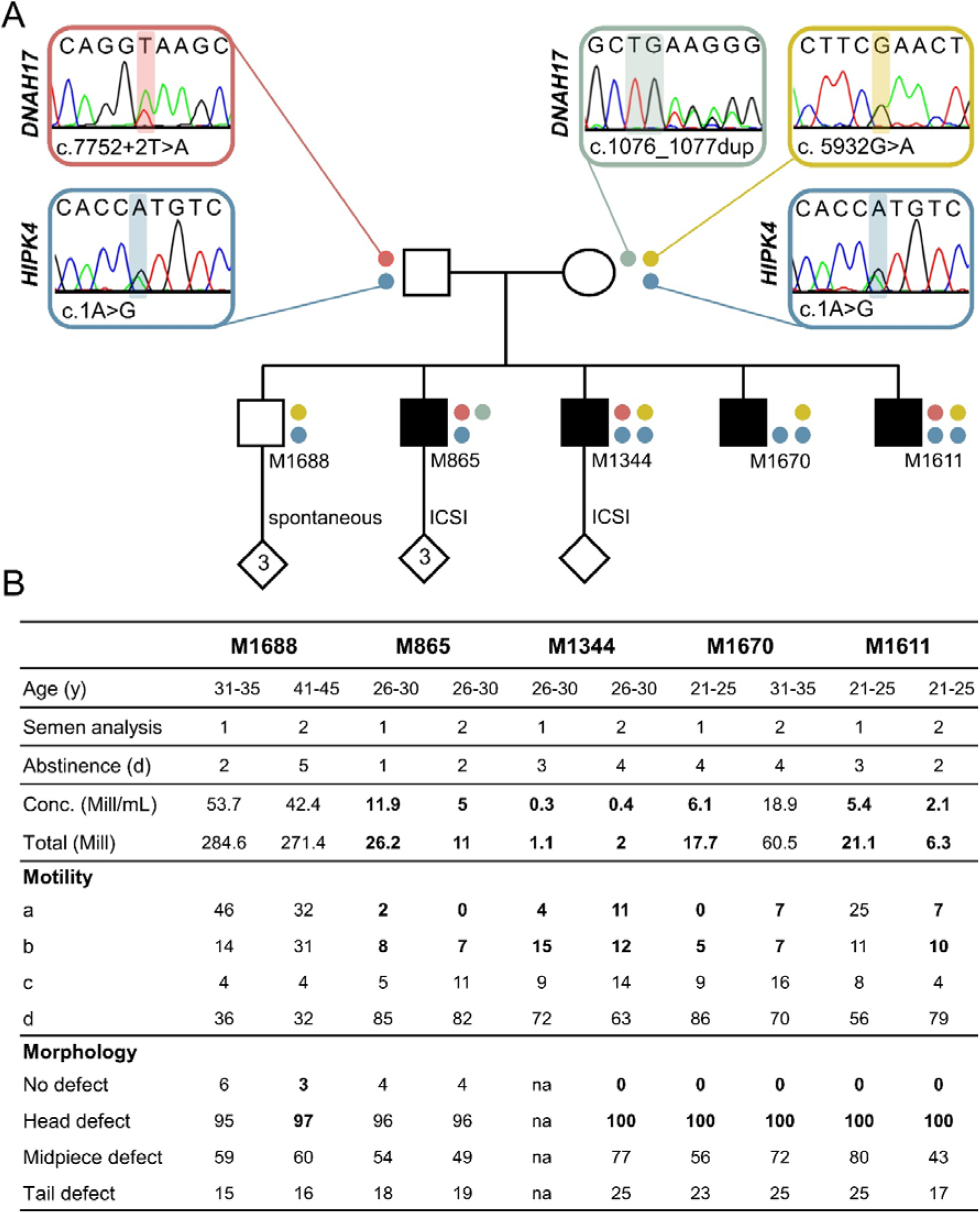
Pedigree, segregation of *HIPK4* and *DNAH17* variants, and semen phenotypes. (A) Filled symbols depict men who are infertile. The coloured dots represent a specific variant in the monoallelic state as indicated by the Sanger traces of the parents. For instance, both parents are heterozygous for c.1A>G in *HIPK4* (blue circle), leading to heterozygosity (one blue circle) or homozygosity (two blue circles) in three of their sons. In *DNAH17*, one paternal (c.7752+2T>A p.?) and two maternal (c.1076_1077dup p.(Lys360*) and c.5932G>A p.(Glu1978Lys)) variants are highlighted in red, blue-green and yellow, respectively. (B) The results of the first two complete semen analyses performed at the CeRA according to the WHO guidelines are shown underneath the respective brother. M1344 had a morphology assessment only once (first and third analysis shown). Due to high variability of human semen parameters, all available semen analyses are shown in Supplementary Table S5. Reference values according to WHO 2021: concentration ≥ 16 million/mL; total sperm count ≥39 million; motility: a+b ≥ 30%, a+b+c ≥ 42%; morphology: ≥ 4% normal. Values outside the reference ranges are highlighted in bold. Abbreviations: y: years; na: not available.

First, the exome-based analysis of 32 valid disease genes for qualitative spermatogenic failure and male infertility revealed no shared genetic cause among all four affected men. However, three variants in the autosomal recessive MMAF gene *DNAH17* (NM_173628.4) were identified in different combinations (Fig. 1, Supplementary Fig. S2). M865 is compound heterozygous for two loss-of-function (LoF) variants in *DNAH17*. The variant c.1076_1077dup leads to a premature stop codon in exon 8 of 81 (p.(Lys360*)) while the second variant, c.7752+2T>A p.?, disrupts the canonical splice donor site after exon 49 (SpliceAI score for donor loss: 1.0; Jaganathan *et al*., 2019). Both variants are ultra-rare (gnomAD: 0.0001773 and 0.00003706) and were listed in ClinVar as pathogenic (Variation ID: 3388882) and likely pathogenic (Variation ID: 2582766), respectively. M1344 and M1611 are also heterozygous for c.7752+2T>A p.?, but in a compound heterozygous state with the hitherto undescribed *DNAH17* missense variant c.5932G>A p.(Glu1978Lys). The residue is located within the AAA1 ATPase domain, is conserved among mammals (Supplementary Fig. S3), and has a CADD score of 27.6 (version GRCh37-v1.6). Still, the impact of this amino acid substitution on the protein function remains unclear. In M1670, however, c.5932G>A p.(Glu1978Lys) was the only *DNAH17* variant, identified in a heterozygous state. This means that irrespective of the missense variant’s significance, *DNAH17* variants cannot explain the infertility of all affected brothers.

Next, we performed an exome-wide analysis for variants that are shared among all four affected brothers or among the three without biallelic LoF variants in *DNAH17* (Supplementary Fig. S2). This approach also revealed no shared homozygous, (compound) heterozygous, or hemizygous variants among all four affected brothers (Supplementary Table S6). But the brothers without biallelic LoF variants in *DNAH17* (M1344, M1670, and M1611) are homozygous for a variant identified in *HIPK4* (NM_144685.5). The identified *HIPK4* variant c.1A>G disrupts the canonical start codon which may lead to the use of an alternative downstream start codon or even loss of protein translation. In the fertile brother (M1688), the variant is present in a heterozygous state fitting with the autosomal-recessive inheritance mode for *HIPK4* associated infertility observed in male mice (Crapster *et al*., 2020; Liu *et al*., 2022).

Based on these findings, we first analysed the flagellar ultrastructure associated with the identified variants in *DNAH17*. Next, we investigated the relevance of the *HIPK4* variant using RNA sequencing, *in silico* and *in vitro* analyses, and characterised the sperm morphology associated with *HIPK4* deficiency.

### DNAH17 variants cause an outer dynein arm (ODA) defect and MMAF in M865

We performed TEM of the sperm obtained from M1688 (fertile) and M865, M1344, and M1670 (infertile) to investigate possible ultrastructural alterations associated with the variants identified in *DNAH17*. Axonemal cross sections of all three brothers with the *DNAH17* missense variant in heterozygous (M1688 and M1670) or compound heterozygous state (M1344) exhibited a normal distribution of ODAs in flagellar cross sections (Fig. 2A). In contrast, ODAs were consistently lacking in flagellar cross sections of M865 being compound heterozygous for two LoF variants in *DNAH17* (Fig. 2B). Additionally, we frequently observed axonemal disruption such as missing or supernumerary microtubule doublets in M865’s sample. IF staining of the sperm nuclei and flagella revealed short, bent, or coiled flagella in M865 compared to the control and the fertile brother (Fig. 2C). The observed loss of ODAs and axonemal disruption in sperm of M865 further supports the pathogenicity of the *DNAH17* LoF variants.

**Figure 2.**
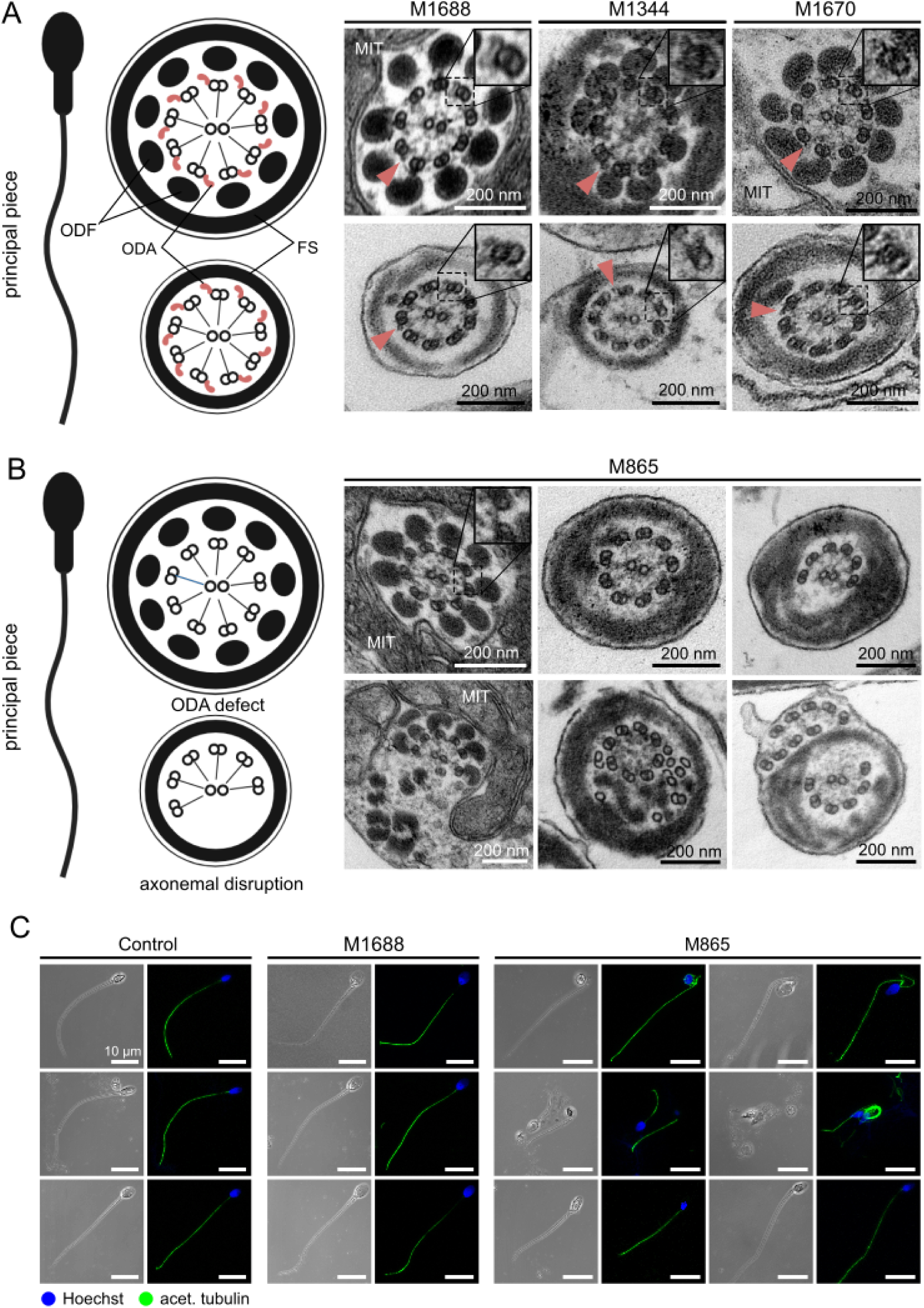
TEM and IF confirming morphological and ultrastructural abnormalities of the sperm flagella in M865 with causal *DNAH17* variants. (A) Left: Schematic representation of regular sperm flagellar cross sections at the proximal and the distal part of the principal piece as seen in transmission electron microscopy (TEM). The axoneme consists of 9 microtubule doublets and a central pair of microtubules. Radial spokes point to the centre, while outer (ODAs, red) and inner dynein arms (not shown) enable flagellar beating. In the proximal principal piece, outer dense fibres (ODFs) and the fibrous sheath (FS) surround the axoneme. Along the principal piece, the ODFs become smaller and fewer until they finally disappear in the distal part. In the midpiece (not shown schematically), the mitochondrial sheath (MIT) replaces the fibrous sheath. Right: TEM images of axonemal cross sections of sperm from M1688, M1344, and M1670. Arrows exemplarily point at ODAs. Note that the upper panel demonstrates flagellar cross sections of M1688 and M1670, which derive from the midpiece as indicated by surrounding MIT, while both pictures from M1344’s sperm represent the principal piece. (B) Left: Schematic representation of a sperm with axonemal disruption associated with *DNAH17* deficiency. Right: TEM images of sperm from M865 clearly show an absence of ODAs as well as axonemal disruption including missing or supernumerary, and misplaced microtubule doublets. (C) Immunofluorescence staining of the nucleus (Hoechst, blue) and the flagellum (acetylated tubulin, green) of control, M1688‘s and M865‘s sperm. Scale bar: 10 µm. Schematic representations in (A) and (B) were created in BioRender (https://BioRender.com/frrmysv).

### HIPK4 is expressed during later stages of spermatogenesis

Data from bulk RNA sequencing of testicular tissue with spermatogenic arrest at different stages indicated that *HIPK4* is neither transcribed in somatic cells nor early stages of human spermatogenesis, but is almost exclusively expressed by samples containing germ cells from the spermatid stage onwards (Fig. 3A). Higher granularity was achieved by single cell-RNA sequencing analysis of testicular tissues with complete spermatogenesis. This revealed that *HIPK4* expression starts in spermatocytes at the end of meiosis 1 (Fig. 3B, Fig. 3C). Based on its expression profile, *HIPK4* is a suitable candidate gene for male infertility due to disturbances in later stages of spermatogenesis.

**Figure 3.**
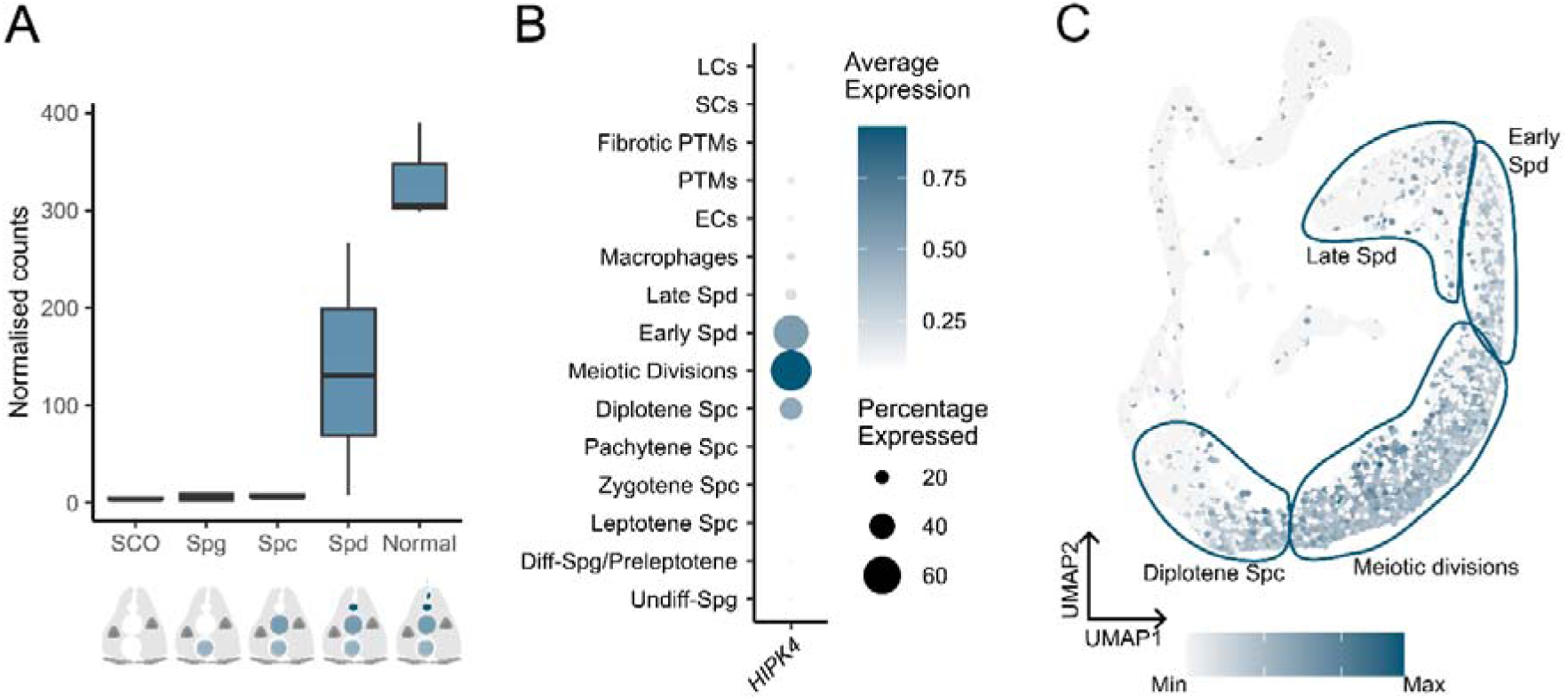
*HIPK4* expression in human testicular tissue. (A) *HIPK4* expression in bulk RNAseq data (Siebert-Kuss *et al*., 2023) from samples without germ cells (Sertoli cell-only), maturation arrest at the spermatogonial (Spg), spermatocyte (Spc), or spermatid (Spd) stage, and full spermatogenesis (Normal). Expression is shown as normalised counts. (B) Dot plot of *HIPK4* expression data derived from single cell RNA sequencing (Di Persio *et al*., 2021) in normal testicular tissue. The size of the circle represents the percentage of cells expressing *HIPK4*, while the intensity of the colour shows the average *HIPK4* expression in each cell type. LCs: Leydig cells; SCs: Sertoli cells; PTMs: peritubular myoid cells; ECs: endothelial cells; Spd: spermatids; Spc: spermatocytes; Diff-Spg: differentiating spermatogonia; Undiff-Spg: undifferentiated spermatogonia. (C) Feature plot of testicular cells expressing *HIPK4* based on the scRNAseq data shown in (B). The level of expression is represented by the intensity of the colour.

### HIPK4 variant leads to a less stable, truncated protein lacking functionally relevant residues

The variant *HIPK4* c.1A>G affects the canonical start codon in transcript NM_144685.5. However, further downstream ATG-codons are located either in frame or out of frame within the first exon of *HIPK4* (Fig. 4A). To analyse the translation initiation *in vitro*, we transfected HEK293T cells with a shortened construct containing WT or mutant *HIPK4* exon 1 (Fig. 4B). Given the difference in protein sizes, protein translation must have initiated at the ATG codon in the mutant sample corresponding to p.Met36 in the WT. Taken together, these data demonstrated that *HIPK4* c.1A>G disrupts the canonical start codon and promotes aberrant translation initiation *in vitro* leading to p.(Met1_Glu35del).

**Figure 4.**
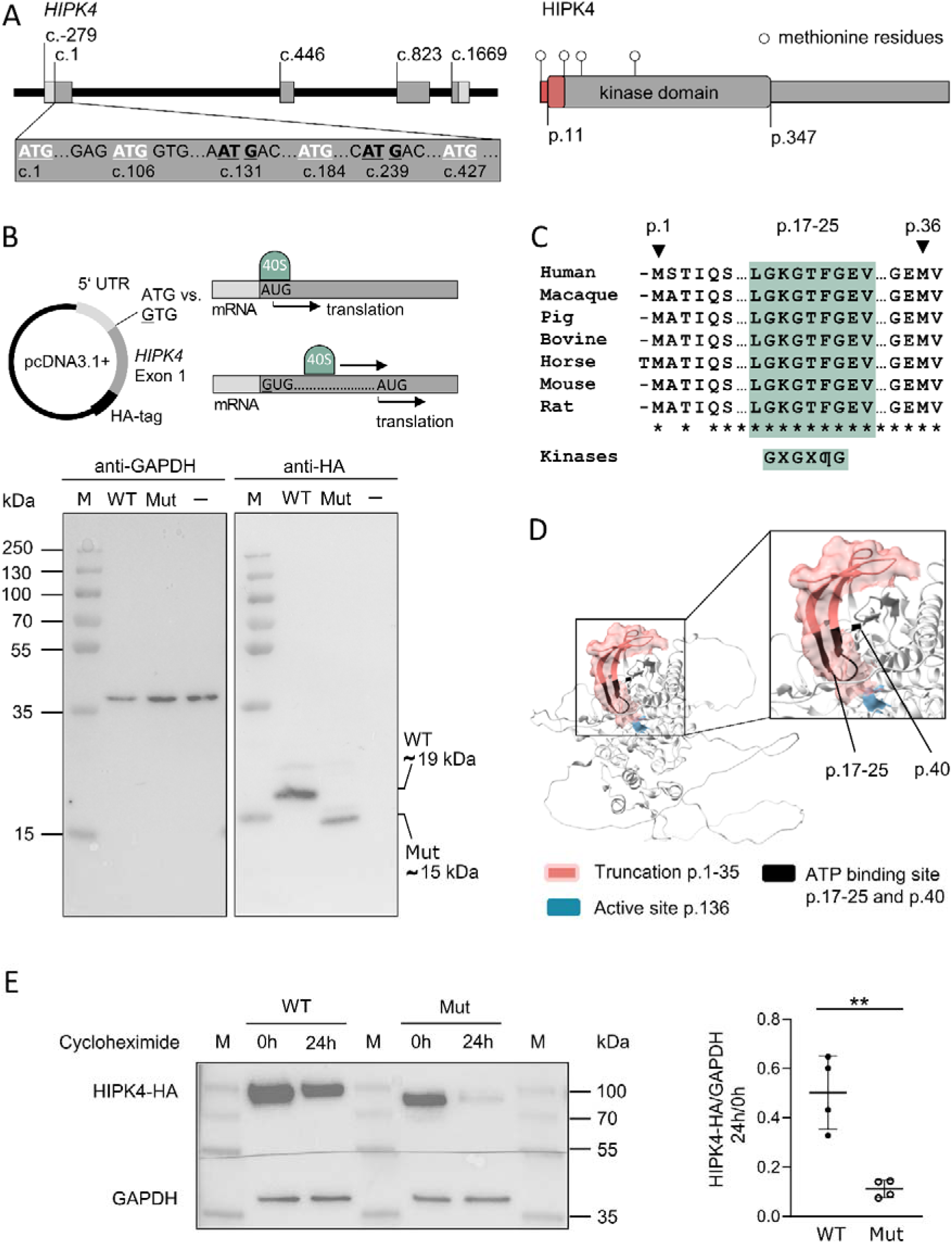
*HIPK4* c.1A>G variant validation. (A) Left: *HIPK4* gene structure with exons in grey, intronic sequence in black, and 5‘ and 3‘ untranslated regions (UTR) in light grey. The position of additional in frame (white) or out of frame ATG triplets (black, underlined) within the first exon is depicted in a close up. Right: Linear protein model of HIPK4 containing a kinase domain (box). White pins indicate the position of methionine residues encoded by in frame AUG. The first 35 amino acids upstream of the second methionine are highlighted in red. (B) Heterologous expression of a construct containing *HIPK4* exon 1 (grey) flanked by its endogenous 5‘ UTR (light grey) and a C-terminal HA-tag (black) in HEK293T cells to test protein translation of the wild type (WT) versus the mutant (Mut) construct. A representative Western blot image (n = 4) of WT, Mut and untransfected cells (−) is shown in relation to a protein marker (M). The observed sizes of the protein fragments match the expected molecular weight for the WT (18.2 kDa) and for the mutant (14.3 kDa) sample if translation initiates at the first in frame AUG codon (p.Met36). (C) Multiple sequence amino acid alignment of HIPK4 orthologues from different mammalian species. Less than 65% of the sequence of human HIPK4 (ENSG00000160396) matched the 1:1 orthologue of all 14 fish and 20 bird/reptile species listed by Ensembl and vice versa. *Drosophila melanogaster* and zebrafish do not have a 1:1 ortholog of human HIPK4. Sequences were retrieved from Uniprot. Stars indicate conserved residues. The phosphate-binding loop (p.17-25, green) contains a Glycin-rich sequence motif present in all typical protein kinases (Röhm *et al*., 2021) with φ being a hydrophobic amino acid like phenylalanine in case of HIPK4. (D) 3D AlphaFold model of HIPK4, adapted in ChimeraX. The N-terminal truncation (red) also encompasses p.17-25 (black) directly interacting with the ATP‘s phosphate (Uniprot Q8NE63). The functionally indispensable residues p.Lys40 and p.Asp136 (He *et al*., 2010) are highlighted in black and blue, respectively. (E) Cycloheximide chase experiment of WT and Mut HIPK4 in transiently transfected HEK293T cells. Cells were either untreated or incubated with cycloheximide for 24 h. GAPDH served as internal control of cellular protein expression. The experiment was carried out four times independently. The graph displays normalised HIPK4 levels (HIPK4-HA/GAPDH) after 24 h relative to its amount at 0 h. The proportion of residual protein amounts after 24 h is significantly lower in the mutant sample compared to WT (paired t-test with significance level of p < 0.05).** p < 0.01.

The alternative translation initiation site used in the mutant (p.Met36 in the WT) is located within HIPK4’s kinase domain (Fig. 4A). The part of the domain affected by the truncation is conserved between mammalian species but not in lower eukaryotes and shares similarity with the catalytical domains of other protein kinases (Fig. 4C). Residues p.17-25 form one part of the ATP binding pocket that is complemented by p.Lys40 (Fig. 4D). To test the mutant protein’s stability, HEK293T cells were transfected with full-length WT and mutant *HIPK4* cDNA followed by translation inhibition with cycloheximide to analyse protein degradation within a 24 h period (Fig. 4E; Supplementary Fig. S4). Protein amounts of both the WT and the mutant decreased over time, but the relative amount of HIPK4 present at the end of the experiment (24 h/0 h) was significantly lower in the mutant than in the WT sample (p = 0.008). These data indicate that the N-terminal truncation results in impaired HIPK4 stability *in vitro*.

### Variable sperm head abnormalities are associated with loss of HIPK4 function

Modified Papanicolaou staining of the fertile brother’s (M1688) ejaculate revealed normally shaped sperm in line with the diagnostic semen analysis (Fig. 5). In contrast, M1670’s sperm showed variable morphological abnormalities like tapered, round, small, and amorphous sperm heads, heads with small or completely absent acrosome, as well as headless sperm tails (acephalic sperm or “pinheads”). Besides, the staining showed abnormalities of the midpiece and flagellum like bent or enlarged midpieces and sperm tails coiled around the head. IF staining of the sperm of all three brothers homozygous for the *HIPK4* variant (M1344, M1670, M1611) was performed to combine the sperm shapes in bright field microscopy with the signal of its nuclei (Hoechst 33342) and flagella (acetylated alpha-tubulin). In line with the Papanicolaou staining of M1670, the additional stainings showed variable sperm head shapes and sizes, as well as short and coiled flagella (Fig. 5C).

**Figure 5.**
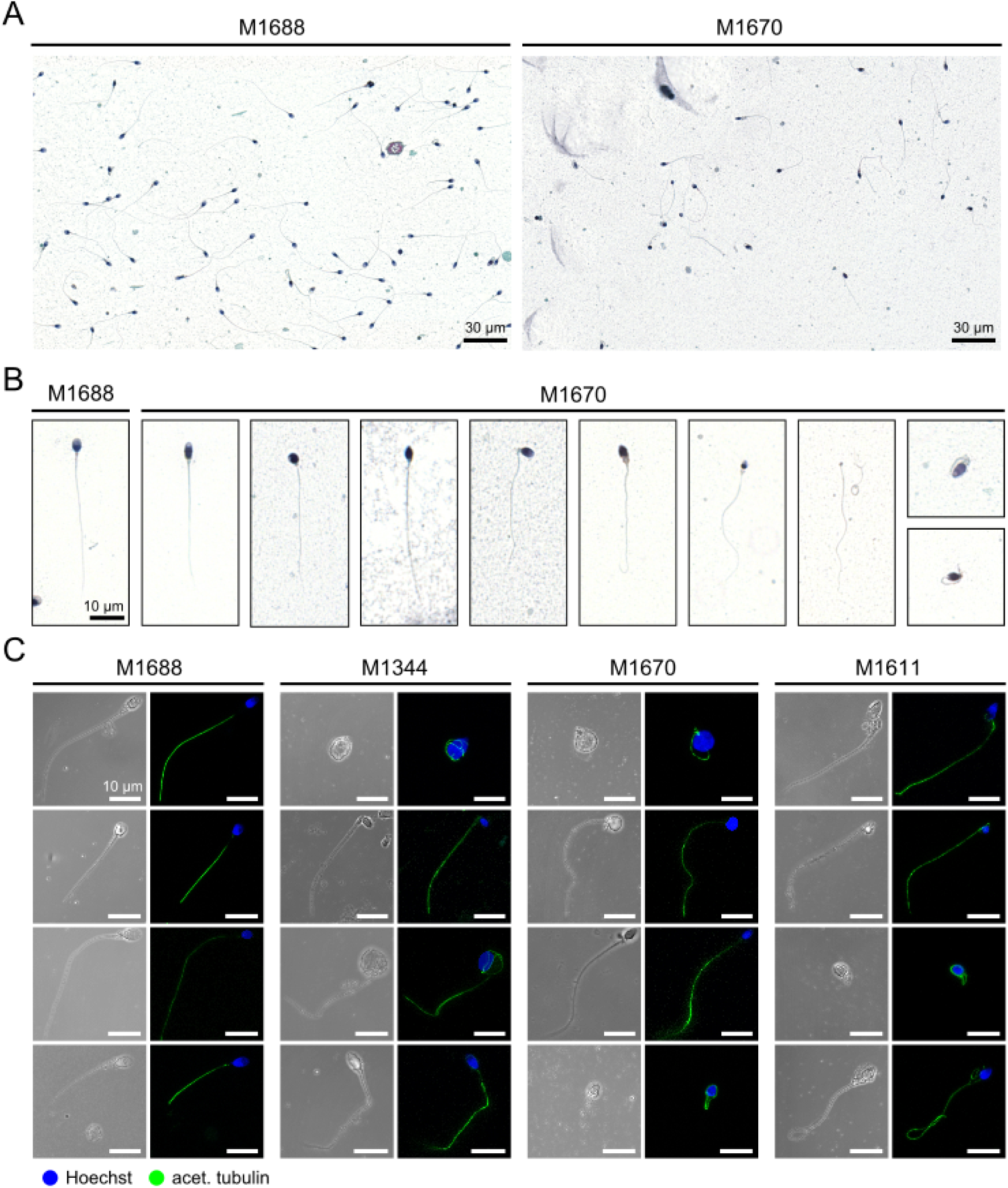
The homozygous *HIPK4* variant is associated with variable sperm head abnormalities. (A) Overview of sperm smears stained with modified Papanicolaou from the fertile brother (M1688) and one of the brothers affected by infertility (M1670), who are heterozygous and homozygous for c.1A>G p.(Met1_Glu35del) in *HIPK4*, respectively. (B) Close-up of a morphologically normal sperm of M1688 and examples of different morphological abnormalities of sperm heads, midpieces, and flagella in M1670’s semen sample. From left to right: tapered, round, amorphous sperm head, bent midpiece, amorphous head and enlarged midpiece, small and round head, headless sperm flagella, and short flagella coiled around the head. (C) Immunofluorescence (IF) staining of sperm from all three brothers homozygous for c.1A>G p.(Met1_Glu35del) in *HIPK4* (M1344, M1670, M1611) compared to the fertile brother (M1688) being heterozygous. The nucleus is shown in blue colour (Hoechst), the flagellum in green (acetylated tubulin). Scale bar: 10 µm.

### Gene and variant assessment according to clinical guidelines

*DNAH17* has already reached a definitive level of evidence for its gene-disease relationship with asthenoteratozoospermia/MMAF (Stallmeyer *et al*., 2025). For *HIPK4*, we collected and scored available experimental and genetic evidence according to ClinGen. To establish the gene-disease relationship, it is mandatory to identify further independent cases with a fitting phenotype. So far, no other patient with biallelic *HIPK4* variants and teratozoospermia has been reported. A published homozygous *HIPK4* missense variant identified in a man with azoospermia was not considered due to uncertain significance and the different phenotype (Alhathal *et al*., 2020; Supplementary Table S7, Supplementary Table S8). By analysis of exome/genome data of men with infertility and different semen phenotypes including crypto-/azoospermia as well as oligo-/astheno-/teratozoospermia in the MERGE cohort, we did not identify another man with biallelic high-impact variants. Therefore, the gene-disease relationship cannot be formally established yet despite a preliminary score of 7 points which is equivalent to a moderate level of evidence (Supplementary Table S7). Thus, *HIPK4* remains a promising candidate gene for human teratozoospermia with convincing experimental but so far sparse genetic evidence. According to the ACMG guidelines, both *DNAH17* LoF variants (c.1076_1077dup p.(Lys360*) and c.7752+2T>A p.?) were classified as pathogenic and the *DNAH17* missense variant (c.5932G>A p.(Glu1978Lys)) as variant of uncertain significance (VUS). Assuming a valid gene-disease relationship for *HIPK4*, the variant c.1A>G p.(1Met_35Gludel) in *HIPK4* was classified as likely pathogenic (Supplementary Table S7).

## Discussion

Within the same family, we discovered biallelic variants in *DNAH17* and *HIPK4* likely causing qualitatively impaired spermatogenesis in four brothers. While M865’s infertility is caused by compound heterozygous LoF variants in *DNAH17* leading to an ODA defect and MMAF, we present data that the homozygous *HIPK4* variant c.1A>G p.(1Met_35Gludel) explains the infertility of his three brothers. Via heterologous expression in HEK293T cells, we demonstrate that the mutant protein is truncated lacking a part of its kinase domain and is less stable. Matching HIPK4’s expression in later stages of spermatogenesis, all three affected brothers presented with teratozoospermia and sperm head defects most frequently accompanied by oligo- and asthenozoospermia.

The occurrence of two distinct genetic causes leading to an apparently similar condition might be surprising. But, given the considerable heterogeneity of male infertility already regarding validated genes (Stallmeyer *et al*., 2025) and moreover the sheer number of genes expressed in the testis (Uhlén *et al*., 2015), this is not expected to be a unique phenomenon. Further, consanguinity of the parents increases the probability of homozygosity of (likely) pathogenic variants. Thus, exome analysis of affected and unaffected family members, combined with detailed phenotypic characterisation, is advantageous in confirming or rejecting the hypothesis of a shared genetic basis, and in providing better counselling concerning the recurrence risk and treatment options.

M865’s phenotype is in line with previous reports of *DNAH17* LoF or missense variants causing asthenozoospermia and MMAF (see Song *et al*., 2023 for a review). Loss of DNAH17 function in M865 results in a consistent absence of ODAs with largely preserved 9+2 axonemal organisation, while axonemal disruption represents an additional, variable feature. The severely reduced progressive sperm motility observed in M865 and in previously reported patients (Song *et al*., 2023) is consistent with the functional consequences of ODA loss described by Whitfield *et al*. (2019).

Concerning *HIPK4*, human variants have been previously published in the context of azoospermia (Alhathal *et al*., 2020; Liu *et al*., 2022). However, the semen analysis of all three men homozygous for the *HIPK4* variant presented here revealed teratozoospermia which is in line with the published mouse models (Crapster *et al*., 2020; Liu *et al*., 2022). Like in *Hipk4*^−/−^ mice, unbiased semen analyses that were performed prior to genetic testing showed 100% sperm head defects in all three brothers affected by the homozygous start-loss variant. Morphological sperm analyses revealed variable head abnormalities instead of monomorphic teratozoospermia. Although we cannot rule out a phenotypic spectrum ranging from severe teratozoospermia (with or without oligo- and asthenozoospermia) to azoospermia, our data indicates that loss of *HIPK4* function is compatible with completion of spermatogenesis in humans and extrapolating from three affected brothers makes a phenotype of azoospermia as a result of HIPK4 deficiency unlikely.

Importantly, we clarified the inheritance mode as autosomal recessive. Unlike Liu *et al*. (2022), presenting solely heterozygous *HIPK4* variants, all three affected men in this family are homozygous for c.1A>G p.(1Met_35Gludel) in *HIPK4*. As both the father and the fertile brother are heterozygous for the same variant, we provide evidence that *HIPK4* is a recessive gene associated with male infertility. This is in line with preserved fertility in *Hipk4*^+/−^ mice producing comparable litter sizes as WT males (Crapster *et al*., 2020). Taken together, these findings argue against a causal association between the heterozygous variants published by Liu *et al*. (2022) and the respective patients’ azoospermia. Alhathal *et al*.(2020), however, identified a homozygous missense variant in *HIPK4* (NM_144685.3:c.935C>T p.(Ala312Val)), but no functional validation was performed, leaving the variant’s significance uncertain.

In contrast, we present a truncating variant in *HIPK4* (c.1A>G p.(1Met_35Gludel)). The mutant HIPK4 protein lacks one part of the ATP binding site (p.17-25), a hydrophobic pocket that is formed at the interface of two lobes (Arter *et al*., 2022; Röhm *et al*., 2021). Only four amino acids remain prior to the lysine residue (p.40) that contributes to ATP binding and is essential for kinase function (He *et al*., 2010). Thus, an N-terminal truncated protein is expected to severely affect the kinase function via impaired ATP binding. Based on our experiments, we cannot rule out a residual function of the truncated HIPK4 protein. Still, the cycloheximide assay indicates reduced protein stability compared to the WT protein *in vitro* arguing against a sufficient function. Despite the limitation that functional analysis of patient-derived cells was not possible because of *HIPK4* expression in the testis, use of HEK293T cells was suitable to analyse the variant’s effect at the protein level assuming that translation initiation is conserved between eukaryotic cells (Kozak, 1999).

Serine/threonine kinase 33 (STK33) is another testis-expressed protein kinase reported to be required for male fertility in mice and men. Ma *et al*. (2021) published a homozygous frameshift variant in *STK33* identified in four infertile brothers with asthenozoospermia and MMAF, which was in line with the murine phenotype (Martins *et al*., 2018). Based on the fact that transcription ceases in elongating spermatids, post-transcriptional and post-translational modifications such as phosphorylation become even more important for the regulation of differentiation and maturation processes (Baker, 2016; Ogurtsov *et al*., 2008). Given the high number of testis-specific protein kinases (Ogurtsov *et al*., 2008), it is likely that further kinases other than HIPK4 or STK33 might be related to male infertility.

Among the three brothers homozygous for the truncating *HIPK4* variant (M1344, M1670, and M1611), semen parameters varied regarding sperm count and motility. A similar intrafamilial variability of semen parameters has also been observed in three infertile brothers homozygous for the same frameshift variant in *STK33* (Ma *et al*., 2021). The semen phenotype might be modified by the individual genetic background as well as by environmental factors. As the clinical significance of the *DNAH17* missense variant remains unclear, we cannot rule out a contribution of the identified *DNAH17* variants to M1344’s and M1611’s semen phenotypes. The fact that sperm motility reached normal values at least once in M1611 and flagellar ultrastructure differed from M865, however, argues against the *DNAH17* missense variant’s significance.

*HIPK4* is primarily expressed in the testis, to a lower extent in the brain, and in low levels in other tissues (Karlsson *et al*., 2021; Uhlén *et al*., 2015). Both the clinical data obtained from our patients and the *Hipk4*^−/−^ mouse model do not suggest any clinical consequences apart from male infertility. Likewise, additional phenotypes were neither observed in patients with a homozygous *STK33* LoF variant nor in *Stk33*^−/−^ mice despite a broader expression profile than expected for a gene causing isolated infertility (Ma *et al*., 2021; Martins *et al*., 2018). As *HIPK4* is also highly expressed in human oocytes, its role in female (in)fertility is yet to be discovered. Female *Hipk4*^−/−^ mice, however, displayed normal fertility (Crapster *et al*., 2020).

Understanding genetic causes of male infertility also enables target identification for contraception. Ku *et al*. (2024) very recently developed a selective STK33 inhibitor as male contraceptive. In mice, inhibition of Stk33 reversibly affected sperm motility and in higher doses also caused teratozoospermia, reproducing the effect of complete knockout. Likewise, *HIPK4* has been proposed as a potential target for contraception (Crapster *et al*., 2020).

Concerning treatment options, both human and murine data indicate that assisted reproduction can be successful in men with biallelic *HIPK4* variants. However, based on reduced oocyte binding of HIPK4-deficient sperm in mice (Crapster *et al*., 2020), it might be advisable to perform ICSI in men with HIPK4-related infertility.

Based on the genetic evidence in this study and published experimental data, *HIPK4* represents a promising candidate gene for male infertility. Alhathal *et al*. (2020) have already evaluated the gene-disease relationship for *HIPK4* and male infertility according to ClinGen. Despite sparse genetic data consisting of a single homozygous missense variant identified in an azoospermic patient and classified as of uncertain significance, they reported a moderate evidence level. According to Strande *et al*. (2017), however, occurrence of a single homozygous variant of uncertain significance should be regarded as providing only limited evidence. This is the first report of *HIPK4* associated with human teratozoospermia (regularly accompanied by oligo- or asthenozoospermia), which is discrepant from the phenotype described by Alhathal *et al*. (2020) and needs further validation in future studies.

## Conclusion

Based on the results in this study, we propose *HIPK4* as candidate gene for human male infertility due to sperm head defects, typically also associated with impaired sperm motility and reduced sperm counts (oligoasthenoteratozoospermia). Importantly, HIPK4 deficiency is compatible with successful medically assisted reproduction via ICSI. Further replication is required to firmly establish the gene-disease relationship and to clarify the phenotypic spectrum. This study also highlights that apparently similar phenotypes of male infertility within a family might have distinct genetic causes.

## Supplementary data

Supplementary data are available at Human Reproduction Open online.

## Data availability

All data underlying this article not included already will be shared on reasonable request to the corresponding author with the exception of the exome sequencing data because of the probands’ privacy protection of genetic data. All genetic variants in *DNAH17* and *HIPK4* have been submitted to ClinVar (SUB15989334).

## Acknowledgments

The authors kindly thank Hubert Schorle and Andjela Kovacevic for their opinions and support, Christina Burhöi, Luisa Meier, Ann-Kristin Dicke, Michelle Diane Runkel, Sironi Sivalingam, Johanna Kuß, Daniela Hanke, and the team of the andrology lab at the CeRA for technical and methodological support, as well as Tzviya Zeev Ben Mordehai for sharing expertise concerning sperm electron microscopy.

## Author contributions

Study conceptualisation: S.A.K., C.F., F.T. Data curation: S.A.K., C.R., C.K., S.L., C.F., B.S., N.N., S.D.P., S.K., H.O. Funding acquisition: S.A.K., A.S.G., R.T.W., N.N., J.R., H.O., S.L., C.F., F.T. Investigation: S.A.K., C.R., I.A., R.T.W., S.D.P., N.N., S.L. Visualisation: S.A.K., C.R., S.L., A.S.G. Writing of original draft: S.A.K., C.F. Review and editing: S.A.K., C.R., I.A., C.K., A.S.G., J.W., R.T.W., S.D.P., N.N., J.R., H.O., S.L, S.K., B.S., C.F., F.T. All authors revised and approved the final version of the manuscript.

