## Supplementary data for "*HIPK4* is a novel gene associated with teratozoospermia and male infertility"

### Table of Content

**Supplementary Tables**

**Supplementary Figures**

### Supplementary Tables

**Supplementary Table S1. Primer sequences.**

| Purpose | Primer sequence (5'-3') |
| --- | --- |
| **Sanger validation of variants** | |
| *HIPK4* c.1A>G | Forward: TCCAGCAGCTCAAAGACCAG; Reverse: TCACTCCTGCCTTATCCCCA |
| *DNAH17* c.1076_1077dup (exon 8) | Forward: TAGGGTAGGAGCAGTCTGGG; Reverse: ATTTATGCTGCAGCCTTGGC |
| *DNAH17* c.5932G>A (exon 39) | Forward: ATGAATACGGGCAGGTCGTC; Reverse: AGGAGGGAAGTAGGGAGCAG |
| *DNAH17* c.7752+2T>A (exon 49) | Forward: CTGGCTAGCTGCTCATCCTC; Reverse: GCAGGAGAATGGGGCGAA |
| **Cloning and mutagenesis** | |
| Cloning of *HIPK4* 5' UTR and exon 1 (complementary sequence in bold) introducing restriction enzyme sites and a premature stop codon (italic) | Forward (HindIII): GGTGGTAAGCTT**ACTCAACAGCGCTGGAACCCATTCGG**<br>Reverse (XhoI): ACCACCCTCGAG*TTA***CTTGACCCTGAAGGGGCAGCGGGTCT** |
| Subsequent cloning of HA-Tag using phosphorylated primers | Forward (HA-tag): TACCCATACGATGTTCCAGATTACGCTTAACTCGAGTCTAGAGGGCCCGTTTAAACC<br>Reverse: CTTGACCCTGAAGGGGCAGCGGGT |
| PCR and sequencing of the insert | Forward: GTAACAACTCCGCCCCATTG<br>Reverse: AGGAAAGGACAGTGGGAGTG |
| Cloning of HIPK4 5' UTR and complete cDNA | Forward (HindIII): GGTGGTAAGCTT**ACTCAACAGCGCTGGAACCCATTCGG** |

| | |
|---|---|
| | Reverse (XhoI): ACCACCCTCGAG**TCAGTGGTGCCCGGTGACATGCT** |
| Subsequent cloning of HA-Tag using phosphorylated primers | Forward (HA-tag): TACCCATACGATGTTCCAGATTACGCTTGACTCGAGTCTAGAGGGCCCGTTTAAACC<br>Reverse: CTCCAGCATGTCACCGGGCACCAC |
| Mutagenesis c.1A>G | Forward: ACCGTGTCCACCATCCAGTCGGAGACTGA<br>Reverse: TCAGTCTCCGACTGGATGGTGGACACGGT |
| Sequencing of the HIPK4 construct | Insert primer 1: GTAACAACTCCGCCCCATTG<br>Insert primer 2: CTTCAGCAACCTCATTCGGC<br>Insert primer 3: AGGAAAGGACAGTGGGAGTG<br>Standard pcDNA3.1(+) primer:<br>AmpStart: AAAACAGGAAGGCAAAATGC<br>AmpStop: TCAGGCAACTATGGATGAAC<br>BGH Reverse: TAGAAGGCACAGTCGAGG<br>CMV Forward: CGCAAATGGGCGGTAGGCGTG<br>pCI Reverse: GCAATAGCATCACAAATTTCAC |

Abbreviations: UTR: Untranslated region

**Supplementary Table S2. Antibody information.**

| Antibody | Manufacturer | Identifier/Catalogue number | Dilution |
|---|---|---|---|
| **Immunofluorescence staining** | | | |
| **Primary antibody** | | | |
| Mouse anti-acetylated alpha-tubulin | Sigma-Aldrich, Germany | T6793 | 1:4,000 in 1% BSA in PBS |
| **Secondary antibody** | | | |
| Goat anti-mouse Alexa Fluor 488® | Thermo Fischer Scientific, USA | A11029 | 1:1,000 1% BSA in PBS |
| **Western Blot** | | | |
| **Primary antibody** | | | |
| Rat anti-HA-tag | Roche (Merck, Germany) | 11867423 | 1:2,000 in 5% milk powder in TBST |
| Rabbit anti-GAPDH | Cell Signaling Technology, USA | 5174 | 1:1,500 in 5% milk powder in TBST |
| **Secondary antibody** | | | |
| Goat anti-rat | Sigma-Aldrich, Germany | A9037 | 1:1,000 in 5% milk powder in TBST |
| Mouse anti-rabbit | Santa Cruz Biotechnology, USA | 2357 | 1:1,000 in 5% milk powder in TBST |

Abbreviations: BSA: Bovine serum albumin; PBS: Phosphate Buffered Saline; TBST: Tris-buffered saline with Tween20

32 **Supplementary Table S3. Amino acid sequences and functional annotation.**

**Amino acid sequences (UniProt)**

| **Protein** | **Species** | **UniProt Identifier** |
|---|---|---|
| **HIPK4** | Human | Q8NE63 |
| | Macaque | Q8WP28 |
| | Mouse | Q3V016 |
| | Rat | Q4V793 |
| | Bovine | E1BB49 |
| | Pig | I3LVJ2 |
| | Horse | F6YYR8 |
| **DNAH17** | Human | Q9UFH2 |
| | Macaque | A0A2K6DCQ0 |
| | Mouse | Q69Z23 |
| | Rat | D4A599 |
| | Bovine | E1BLB4 |
| | Pig | A0A8W4FH26 |
| | Horse | F7BXB8 |

| | | |
|---|---|---|
| | Platypus | F6SCH9 |
| **Functional annotation** | | |
| **Protein (UniProt ID)** | **Domain or binding site** | **Amino acids** |
| HIPK4 (UniProt: Q8NE63) | Protein kinase domain | 11-347 |
| | ATP binding site | 17-25 and 40 |
| | Active site | 136 |
| DNAH17 (UniProt: Q9UFH2) | ATPases associated with diverse cellular activities (AAA) domain 1 | 1809-2030 |
| | ATP binding site | 1847-1854 |

Abbreviations: ATP: Adenosine triphosphate

**Supplementary Table S4. Rules for assignment of specific ACMG criteria.**

| ACMG criterion | Description |
|---|---|
| **PVS1** | PVS1 was assigned for variants leading to a premature stop codon (PMC) such as nonsense and frameshift variants that are expected to cause nonsense mediated decay (PMC > 50–55 nucleotides from the last exon-intron-junction) if loss-of function (LoF) is a known mechanism or for variants disrupting a canonical splice site (+/-1,2) likely leading to LoF. LoF was assumed for *HIPK4* based on published mouse models (Crapster *et al*., 2020; Liu *et al*., 2022). |
| **PS3** | Functional evidence supporting a deleterious effect led to the assignment of PS3 with one point per experiment (*in vitro* analysis of translation initiation and assessment of protein stability counted as one piece of evidence: PS3_supporting). |
| **PM2** | PM2 was given for variants with MAF < 1% in gnomAD (v4.1.0). |
| **PM3** | PM3 was assigned for variants *in trans* with a (likely) pathogenic variant in the same gene and downgraded to supporting in case of homozygosity or a potential alternative genetic cause that might explain the patient's condition instead. |
| **PM4** | Variants leading to an in-frame deletion or insertion on protein level received PM4. |
| **PP1** | PP1 was not assigned in this study due to identification of two different genetic causes rather than one shared among all affected individuals. |
| **PP3** | PP3 was assigned for missense variants with a CADD ≥20. |

Based on the strength of evidence the corresponding points per criterion were added (very strong: 8; strong: 4; moderate: 2; supporting; 1) resulting in classification as pathogenic (≥ 10), likely pathogenic (6-9) or variants of uncertain significance (VUS, 0-5) (Tavtigian *et al*., 2020).

**Supplementary Table S5. Clinical data including comprehensive semen analysis results.**

Please see separate Excel-file.

39 **Supplementary Table S6. Explorative analysis of variants shared by the affected brothers.**

| Gene (transcript) | Exon | Variant | CADD | MAF gnomAD v2.1.1 | Expression (GTEx) | Comments |
|---|---|---|---|---|---|---|
| **Homozygous variants shared by all four affected brothers** | | | | | | |
| none | | | | | | |
| **Homozygous variants shared by M1344, M1670, and M1611** | | | | | | |
| *PRODH2* (NM_021232.1) | 11/11 | c.1522G>T p.(Val508Leu) | 24.4 | 0.000184 | Not in the testis | Excluded |
| *MAP4K1* (NM_001042600.3) | 18/31 | c.1214C>T p.(Ser405Phe) | 26.5 | 0.00003393 | Not in the testis | Excluded |
| *HIPK4* (NM_144685.5) | 1/4 | c.1A>G p.(Met1?) changed to p.(Met1_Glu35del) | 20.5 | - | Highest in testis, also in the brain | Published mouse models with male infertility and oligoasthenoteratozoospermia (PMID: 32163033; PMID: 35931115) |
| **Compound-heterozygous variants shared by all four affected brothers** | | | | | | |
| none | | | | | | |
| **Compound-heterozygous variants shared by M1344, M1670, and M1611** | | | | | | |
| none | | | | | | |
| **Heterozygous variants (MAF gnomAD < 0.001) shared by all four affected brothers** | | | | | | |
| none | | | | | | |
| **Heterozygous variants (MAF gnomAD < 0.001) shared by M1344, M1670, and M1611** | | | | | | |

| | | | | | | |
|---|---|---|---|---|---|---|
| *EFL1* (NM_024580.6) | 16/20 | c.1849A>C p.(Ile617Leu) | 23.8 | 0.00001203 | Ubiquitous | Autosomal recessive Shwachman-Diamond syndrome 2 (OMIM 617941); excluded |
| *PHOSPHO1* (NM_001143804.1) | 3/3 | c.49_52dup p.(Leu18ArgfsTer353) | 24.2 | - | Testis and blood | *Phospho1*$^{-/-}$ mice have a skeletal phenotype, mating of *Phospho1*$^{+/-}$ mice (and of *Phospho1*$^{-/-}$ mice) produced pubs (PMID: 20684022); no entry in OMIM; no described association with human male infertility; excluded |
| **Hemizygous variants (MAF gnomAD < 0.001) shared by all four affected brothers** | | | | | | |
| none | | | | | | |
| **Hemizygous variants (MAF gnomAD < 0.001) shared by M1344, M1670, and M1611** | | | | | | |
| none | | | | | | |

See Supplementary Fig. S2 for a detailed description of the workflow. Abbreviations: MAF: Minor allele frequency

**Supplementary Table S7. Preliminary ClinGen curation for *HIPK4* as autosomal recessive gene associated with male infertility due to teratozoospermia with or without oligo- or asthenozoospermia.**

| Evidence category | Evidence type | Suggested points | | Points given | PMID/Comments | Total |
|---|---|---|---|---|---|---|
| | | Default | Range | | | |
| Experimental evidence (max. 6 points) | | | | | | 4 |
| Function | Protein Interaction | 0.5 | 0-2 | 0.5 | Interaction with and phosphorylation of RIM-BP3 required for male fertility in mice (Liu et al., 2022; PMID: 35931115) | |
| | Expression | 0.5 | 0-2 | 1 | Expression almost limited to testis (and brain) (Uhlén et al., 2015; PMID: 25613900), single cell expression in spermatids (this study) | |
| Functional alteration | Non-patient cells | 0.5 | 0-1 | 0.5 | Cell shape changes, loss of stress fibres and cytokinesis failure after expression in somatic cells (Crapster et al., 2020; PMID: 32163033) | |
| Models | Non-human model organism | 2 | 0-4 | 2 | Two independent infertile mouse models with OAT (Crapster et al., 2020; PMID: 32163033; Liu et al., 2022; PMID: 35931115) | |
| Genetic evidence (max. 12 points) | | | | | | 3 |
| Variant evidence | Predicted or proven null variant (1.5 points) | 0.5 (per variant) | 0 - 3 (per variant) | 3 | c.1A>G p.(Met1_Glu35del), homozygous, this study | |
| | Other variant type (0.1 points) | 0.1 points (per variant) | 0 - 1.5 points (per variant) | 0 | c.935C>T p.(Ala312Val), homozygous, not counted due to a different phenotype (azoospermia and maturation arrest) and lack of functional validation (Alhathal et al., 2020; PMID: 32719396) | |
| Combined experimental and genetic evidence | | | | | | **7*** |
| | | | | LIMITED | 0.1-6 | |
| | | | | **MODERATE** | **7-11** | |
| | | | | STRONG | 12-18 | |
| | | | | DEFINITIVE | 12-18 + Replicated over time | |

*Due to limited genetic evidence it is impossible to establish the gene-disease relationship despite the preliminary score of 7 points. Identification of at least two more unrelated patients with (likely) pathogenic *HIPK4* variants in homozygous or compound heterozygous state and a similar phenotype is mandatory to introduce *HIPK4* as a diagnostic gene.

Abbreviations: OAT: Oligoasthenoteratozoospermia.

**Supplementary Table S8. ACMG classification of variants.**

| Gene (transcript): variant | ACMG criteria | ACMG class |
|---|---|---|
| **This study** | | |
| *DNAH17* (NM_173628.4): c.1076_1077dup p.(Lys360*) | PVS1 (LoF), PM2 (MAF 0.00003603), PM3 (*in trans* with c.7752+2T>A p.? in M865) | pathogenic |
| *DNAH17* (NM_173628.4): c.7752+2T>A p.? | PVS1, PM2 (MAF 0.0003591), PM3 (*in trans* with c.1076_1077dup p.(Lys360*) in M865) | pathogenic |
| *DNAH17* (NM_173628.4): c.5932G>A p.(Glu1978Lys) | PM2 (MAF 0.00002360), PM3_supporting (*in trans* with c.7752+2T>A p.? in M1344 and M1611 but alternative genetic cause identified in both patients), PP3 (CADD GRCh37-v1.6: 27.6) | VUS |
| *HIPK4* (NM_144685.5): c.1A>G p.(Met1_Glu35del) | PS3_supporting (*in vitro* assays showing truncation and reduced stability), PM2 (absent from gnomAD), PM3_supporting (homozygous), PM4 (inframe deletion N-terminal) | likely pathogenic |
| **Alhathal *et al.*, 2020** | | |
| *HIPK4* (NM_144685.3): c.935C>T p.(Ala312Val) | PM2 (MAF 0.00001302), PM3_supporting (homozygous), PP3 (CADD GRCh37-v1.6: 22.7) | VUS |

Abbreviations: ACMG: American College of Medical Genetics and Genomics; MAF: minor allele frequency (gnomAD, v4); VUS: variant of uncertain significance

### Supplementary Figures

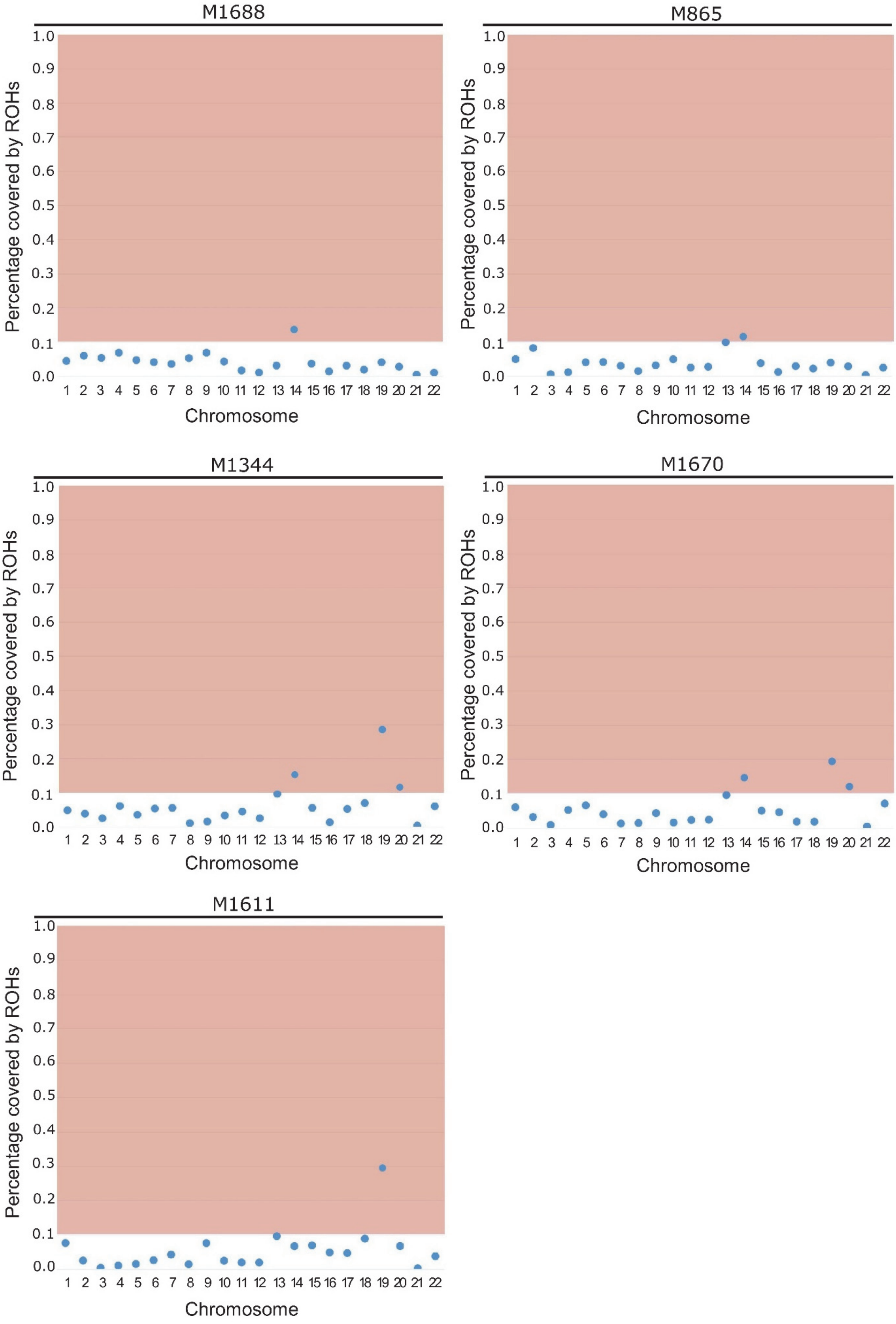

**Supplementary Figure S1. Exome sequencing data shows that the parents are distantly related.**

The exomes of mother, father, and their sons were analysed for regions of homozygosity (ROHs) using the AltAF plotter (Radtke *et al*., 2024). Using the cut-off proposed by the authors (≥ 3 chromosomes with ≥ 0.1 ROH) led to inconclusive results. Exome data of M1344 and M1670 comprises ROH above the threshold. M1344, M1670 and M1611 have increased runs of homozygosity in chromosome 19 including *HIPK4*. The data is compatible with a shared, distant ancestry of the parents.

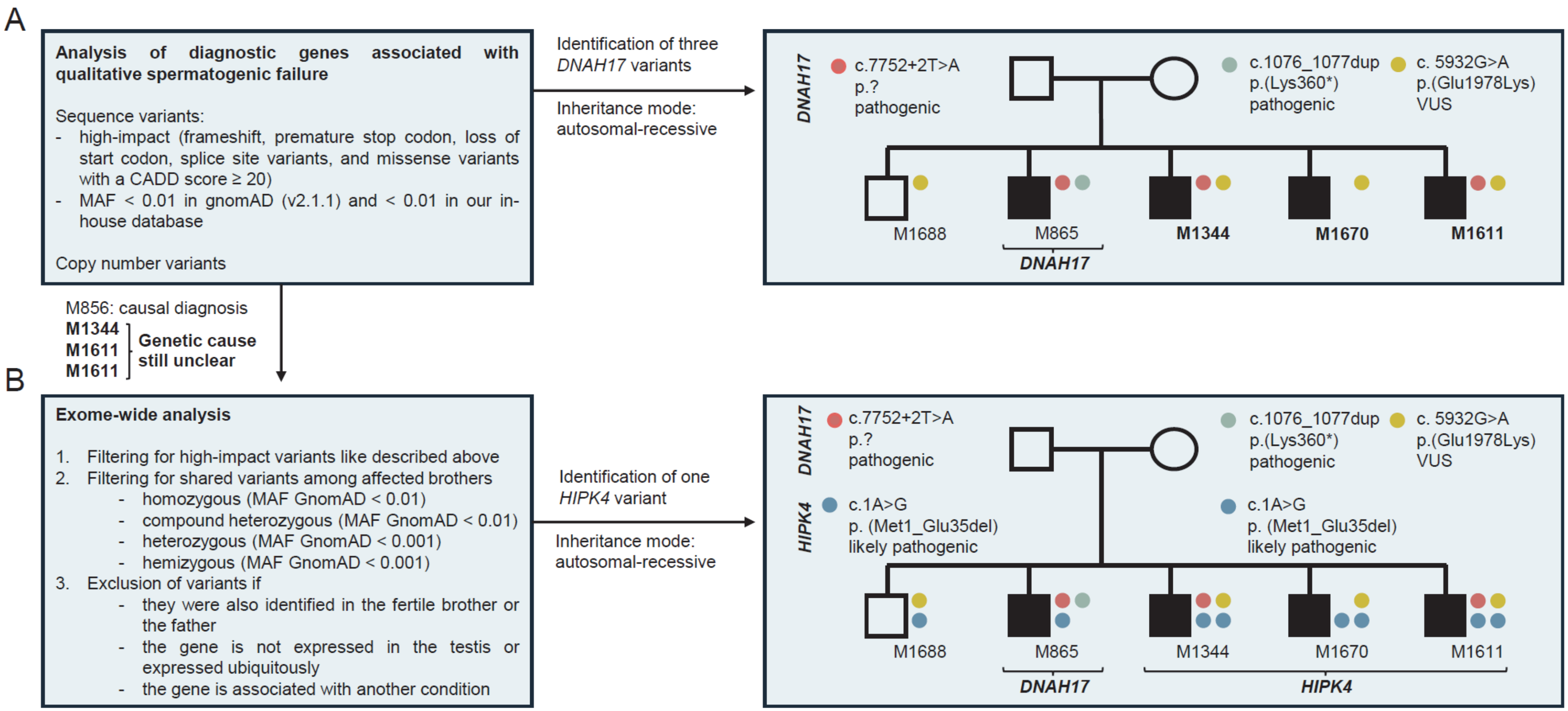


**Supplementary Figure S2. Workflow and main results of the exome-based family analysis.**

(A) 32 genes with an at least moderate level of evidence supporting their gene disease relationship with male infertility and astheno- or teratozoospermia (including other or mixed phenotypes) according to Stallmeyer et al. (2025) were included in the analysis: *ACTL7A, ARMC2, AURKC, CFAP43, CFAP44, CFAP58, CFAP61, CFAP65, CFAP69, CFAP70, CFAP91, CFAP251, DNAH1, DNAH2, DNAH6, DNAH8, DNAH10, DNAH17, DNHD1, DPY19L2, DRC1, FANCM, FSIP2, PLCZ1, PMFBP1, QRICH2, SPAG6, SPEF2, SSX1, SUN5, TSGA10, TTC29*. All analysed genes follow an autosomal recessive (AR) inheritance pattern except for *SSX1* (X-linked). Except for the depicted variants in *DNAH17* (NM_173628.4), no biallelic variants nor hemizygous variants were identified in the affected brothers. Copy number variant (CNV) analysis did not identify any CNV in the analysed genes.

(B) The results of the exome-wide analysis of shared variants are shown in Supplementary Table S6. The homozygous variant in *HIPK4* (NM_144685.5) was identified as the most plausible cause of infertility in M1344, M1670, and M1611.

Abbreviations: MAF: minor allele frequency.

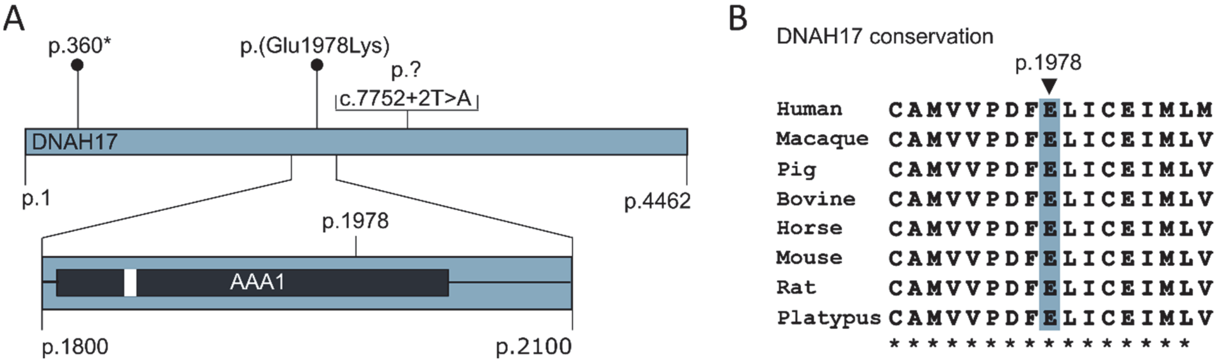


**Supplementary Figure S3. Identified DNAH17 variants on protein level.**

(A) *DNAH17* (NM_173628.4) codes for a protein with 4462 amino acids. The location of identified DNAH17 variants is depicted. Note that c.7752+2T>A disrupts the canonical splice site after exon 49 of 81 and is regarded as loss-of-function variant through aberrant splicing although the exact consequence at protein level is unknown (p.?). The missense variant p.(Glu1978Lys) is located within the first of six 'ATPases associated with diverse cellular activities' (AAA) domains (UniProt: Q9UFH2). The ATP binding site within this domain is marked in white. (B) DNAH17 amino acid sequences from different species were retrieved from UniProt. The Glutamic acid at position 1978 is conserved among mammals.

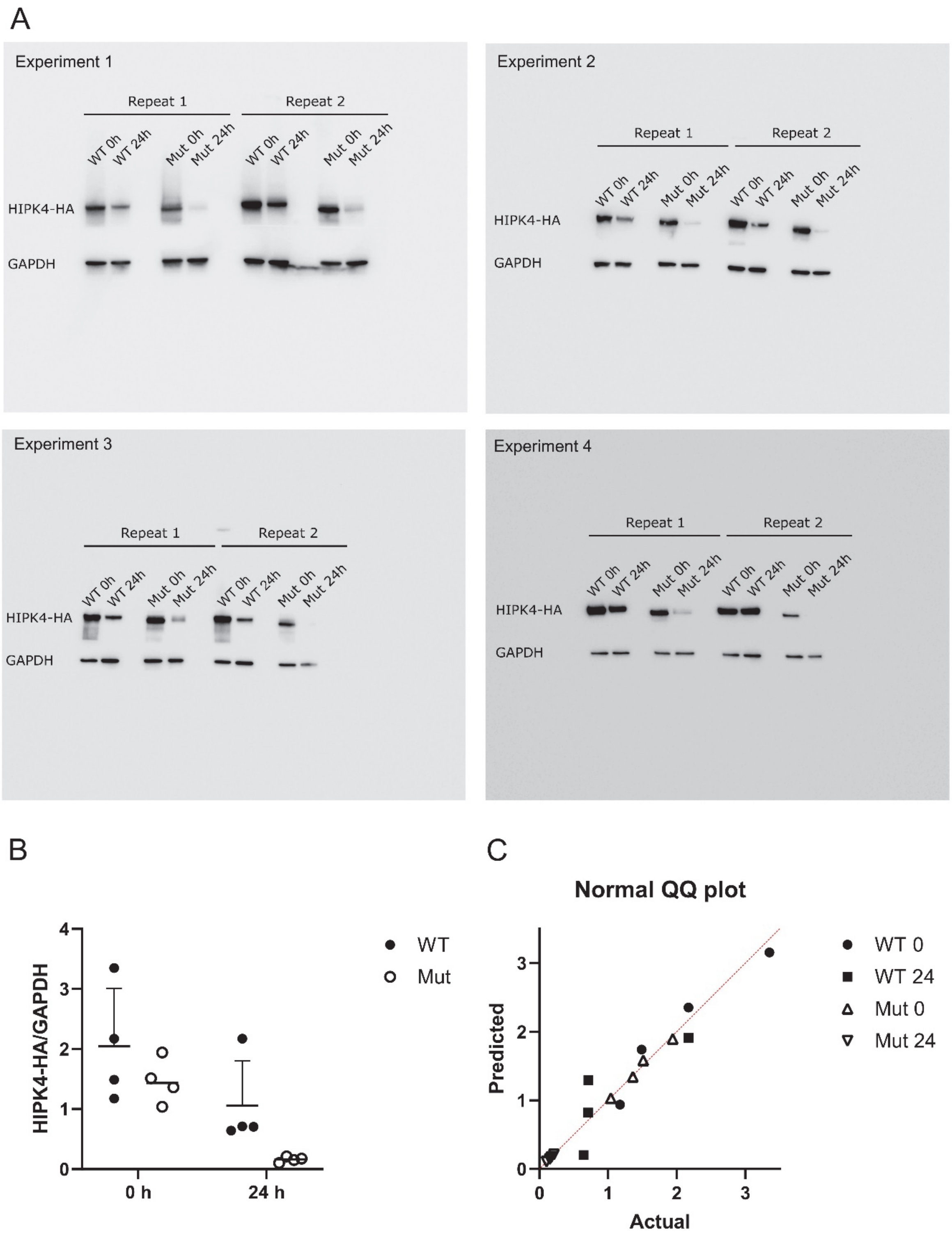


**Supplementary Figure S4. Extended data of the cycloheximide assay.**
(A) The experiment was carried out n = 4 times independently with two technical replicates that were analysed within the same Western blot with imaging of HIPK4-HA and GAPDH simultaneously. Quantification was performed by measuring mean grey values within a rectangular selection used throughout the blot. The background was measured directly above or below the respective band and subtracted. (B) Technical replicates were averaged and

plotted with mean and standard deviation. (C) Visual inspection of the QQ plot was done in advance of the statistical analysis.
